# The public health value of wastewater surveillance for viruses with pandemic potential: a modelling study

**DOI:** 10.64898/2026.09.16.26363050

**Authors:** Amy Dighe, Nicholas C Grassly, Charles Whittaker

## Abstract

Zoonotic spillover and early human-to-human transmission of emerging viruses are often missed by clinical surveillance. Wastewater surveillance (WS) offers low-cost complementary pathogen detection, but its value for emerging viruses remains uncertain. We reviewed evidence of viral shedding in human waste and combined shedding data, detection models and simulated transmission dynamics into a quantitative framework to identify for which types of emerging viruses WS could add most value. Simulations showed that WS improved probability or speed of outbreak detection for viruses shedding >1/100 to >10 times as much as SARS-CoV-2, depending on probability of clinical symptoms and diagnosis. Value added was highest for transmission scenarios with temporally concentrated infections resulting in spikes in daily shedders. Characteristics of SARS-CoV-2, Mpox, Influenza A, Lassa, and Zaire Ebola viruses appear more favourable for WS than others e.g. chikungunya virus. We anticipate this framework can support more targeted, evidence-based application of WS to emerging threats.

## Introduction

Most high-consequence epidemics and pandemics of the past century – including COVID-19, influenza, HIV/AIDS (Human Immunodeficiency Virus/Acquired Immunodeficiency Syndrome), Ebola virus disease and MERS (Middle East Respiratory Syndrome) – have resulted from emergence of novel zoonotic viruses. Zoonotic spillover occurs when ecological conditions, viral traits, and host susceptibility allow viruses circulating in non-human animals to infect humans^1^. While most spillover events do not result in substantial onward transmission, a small fraction are followed by person-to-person spread resulting in outbreaks or, more rarely, pandemics. Land-use change, climate change and globalisation continue to alter ecological systems and increase contact between humans, domestic animals and wildlife, influencing spillover risk and subsequent emergence^2–4^. These trends suggest that zoonotic spillover is likely to remain a key driver of future pandemics.

Despite growing recognition of zoonotic emergence as a driver of pandemics, preparedness efforts remain largely focused on mitigation and control after sustained transmission has already been established in human populations. Pandemics are thought to be preceded by recurring spillover events and stuttering chains of human-to-human transmission^5^, yet the dynamics of these early events remains poorly understood. Clinical surveillance, which depends on healthcare access, symptom severity and diagnostic testing, often fails to detect early transmission. A recent study found that the strongest spatial predictor for outbreak occurrence globally was distance to nearest healthcare facility, highlighting gaps in current surveillance systems and suggesting that many outbreaks go undetected^6^. Even when outbreaks are detected, relying on clinical data alone can lead to delays. Epidemiological evidence suggests that Zaire ebolavirus was circulating in west Africa for months before hospitals clusters led to detection^7^, and initial estimates of outbreak size also suggest a considerable delay in detecting the current Bundibugyo virus outbreak in Democratic Republic of Congo (DRC)^8^.

Wastewater surveillance - the testing of human sewage for pathogens shed by infected individuals - can offer a complementary and comparatively low-cost approach to surveillance that is independent of healthcare access. It has been used for decades to detect poliovirus circulation and remains a core component of eradication efforts, with demonstrable sensitivity^9–11^. More recently, the application of wastewater surveillance for early detection and tracking of SARS-CoV-2 variants of concern^12–14^ has been accompanied by a diversification of approaches to assaying wastewater with varying sensitivity, specificity, and cost, and has driven a rapid expansion in application of wastewater surveillance to other targets, and a growing interest in its use as a biosecurity and pandemic preparedness tool^15^. Beyond early pandemic detection, using wastewater surveillance to reveal patterns of frequent zoonotic spillover and small-scale transmission that currently go undetected could improve understanding of the events preceding pandemics, characterising underlying risk landscape, and informing approaches to pandemic prevention.

However, the public health value of wastewater surveillance is likely to vary substantially across pathogens and epidemiological contexts. Beyond assay performance and wastewater characteristics, detection sensitivity is likely to depend on viral shedding dynamics, transmission dynamics, and surveillance design, yet these factors are rarely evaluated systematically prior to implementation. As a result, it remains unclear for which pathogens wastewater surveillance is most informative, and under what conditions it is likely to add value in terms of increasing detection of frequent zoonotic spillover or outbreaks and gaining valuable response time compared to relying on clinical surveillance alone.

Here, we develop a modelling framework linking viral shedding dynamics, clinical presentation, and transmission dynamics to the performance of wastewater and clinical surveillance for repeated spillover and outbreak detection. Focusing on the 11 viral families identified by the World Health Organisation (WHO) as posing a high risk of a future public health emergency of international concern (a PHEIC)^16^, we synthesise and review available evidence on viral shedding in human waste alongside published data on clinical severity and transmission. We then examine how surveillance performance varies across pathogen profiles and establish a quantitative framework to assess for which types of viruses wastewater surveillance is likely to add the most public health value.

## Results

### Viral shedding to wastewater

We reviewed the existing evidence for viral shedding – specifically sample positivity, amount of virus shed, and temporal shedding dynamics – in human urine and stool for priority viruses from 11 families identified by WHO as well-studied pathogens with available data indicating that they pose a high risk of causing a PHEIC^16^. While we recognise that other biological excreta (e.g. mucus and oral fluids) may enter wastewater, we focus on urine and stool as these likely occur at much higher levels. An existing systematic review was available for shedding of Influenza A Virus (IAV)^17^ and several key studies provided in depth characterisations of the shedding profile of SARS-CoV-2^18–20^. For all other pathogens for which available literature was limited, we conducted a comprehensive literature search and extracted available data (see Supplementary Material for search terms and the extracted dataset (Table S1 & S2)).

The strength of the evidence available to characterise viral shedding varied across pathogens. Strongly informative temporal profiles of viral shedding were only available for SARS-CoV-2 viral copies in stool (urinary shedding is rare^21^) and Mpox virus (MPXV) Ct values in urine^18,19,22^ (Table S3).

Moderately informative quantitative estimates of average viral copies per unit volume of human excreta were available for at least one priority virus in most families^17–19,23–43^ (Figure 1), but reliability was often limited by small sample sizes, infrequent sampling, and bias towards severe cases with access to high-quality healthcare. With the exception of just four studies (focused on SARS-CoV-2 and IAV), the viral load in stool was given in terms of genome copies/ml, and all but one used the wet-weight, which is less comparable than the dry-weight, which would give more accurate measures of genome copies shed per day due to lower variation in production across individuals^20,44^. Viral load estimates varied substantially across families, ranging from 10^3^-10^10^ copies/ml for stool and 10^2^-10^5^ copies/ml for urine. Variation was also evident within families, for example, MERS-CoV estimates were among the lowest, while SARS-CoV-2 were among the highest (Figure 1). Urinary estimates were more commonly available than stool estimates, particularly for *Flaviviridae,* where diagnostic testing often uses urine as viraemia outlasts that in serum^45,46^. Although urinary viral load estimates were available for acute Zika (ZIKV) and dengue virus (DENV) infections, yellow fever virus (YFV) estimated were limited to those based on vaccine strains or convalescent cases^24,28^.

**Figure 1.**
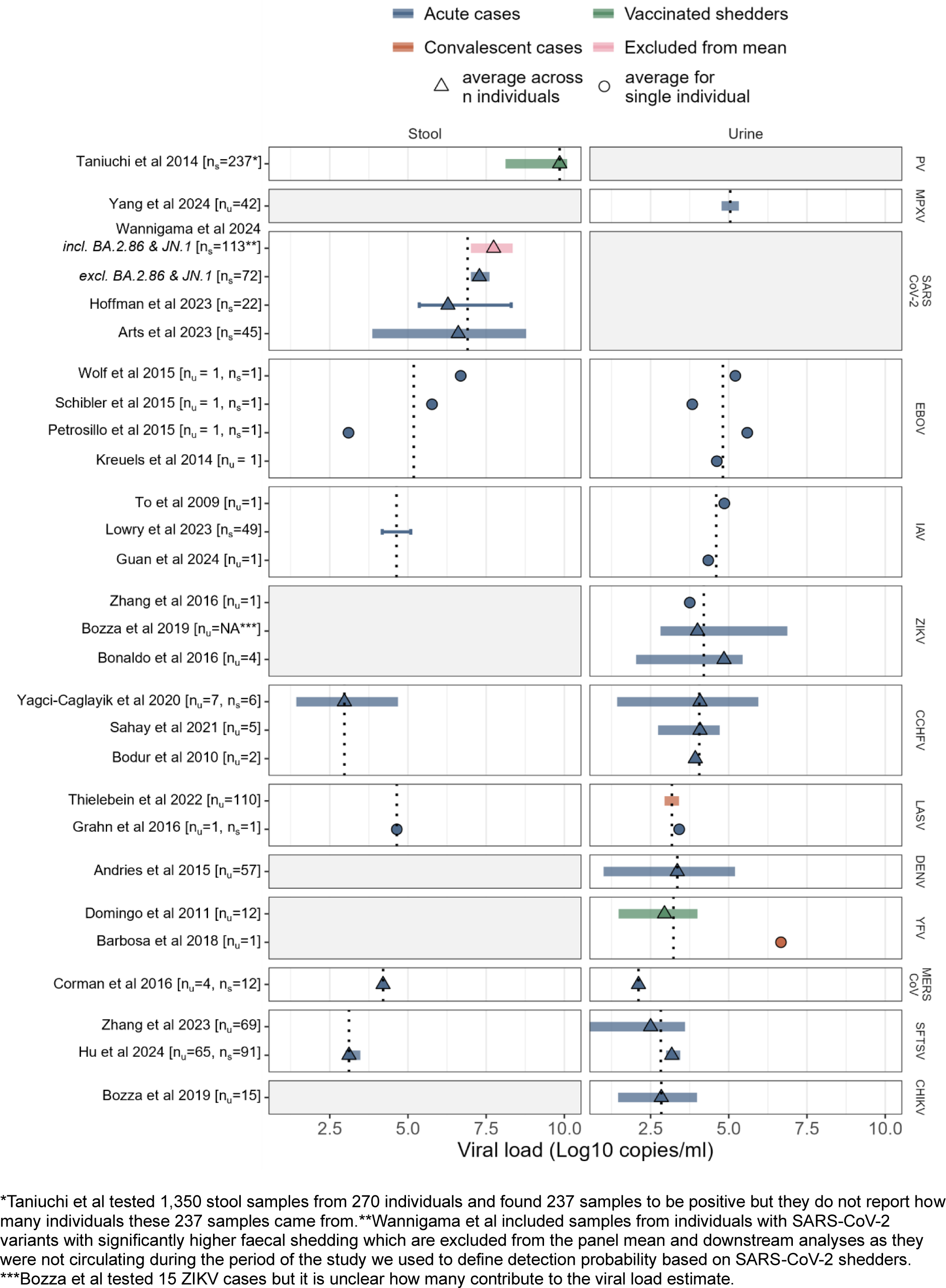
Viral load shed in urine and stool for priority viruses with pandemic potential. From the top, rows are in descending order of total magnitude of daily shedding in stool and urine. Poliovirus (PV) is included for comparison. Estimates are coloured by sampled population sampled: acute cases (blue), convalescent cases (orange), and individuals shedding vaccine-derived virus (green). Thick transparent bars show within-study variability (see Table S1 for the specific variability type represented for each study), and thin error bars show variability or uncertainty estimates from meta-analyses. Vertical dotted lines indicate the sample-size-weighted geometric mean for each virus and sample type on the log scale. Central estimates from single individuals are shown as circles and those from multiple individuals as triangles. All central estimates are means except those from Hu et al 2024 which are medians. In y-axis labels nu and ns denote the number of individuals sampled for urine and stool measures, respectively. Arts et al reported viral load per unit-dry-weight and this was translated to wet weight assuming that an average of 128g wet mass of stool corresponds to a dry mass of 29g^44^. This measure per unit of wet-weight for Arts et al and that reported for Wannigama et al were translated to copies/ml assuming that the density of wet stool is 1.06 g/ml^44^.

Evidence of shedding of priority Paramyxoviruses (Nipah virus, NiV), or the Hantaviruses (Hantaan virus, HTNV and sin nombre virus, SNV) was limited to a small number of weakly informative qualitative reports of the presence of viral genetic material in urine. Four studies detected NiV RNA in urine^47–50^, one detected HTNV RNA in urine from all acute-phase patients tested (4/4)^51^, and another tested urine from convalescent SNV patients sampled after discharge and found them to be negative^52^.

Combining mean viral concentrations in urine and stool with average daily excreta volumes to estimate expected viral load shed to wastewater, MPXV, SARS-CoV-2, Zaire ebolavirus (EBOV) and Influenza A (IAV) exhibited the highest estimated daily loads, ranking below poliovirus. Notably MPXV, SARS-CoV-2 and IAV are already reliably detected through wastewater surveillance. Chikungunya virus (CHIKV), severe fever with thrombocytopenia virus (SFTSV), MERS-CoV and DENV fell into the lower quartile of total expected daily viral load shed to wastewater.

Data on shedding duration were limited by infrequent sampling, and censoring around hospitalisation. Studies reporting duration are presented in Table S4 along with a summary of duration estimates taken forward into downstream modelling. The percentage of patients with a positive sample during acute infection are documented in Table S1 but are likely to represent a considerable underestimate due to infrequent sampling over the infection course.

### Factors affecting the potential public health value added by wastewater surveillance

#### The effect of viral characteristics

We simulated subcritical outbreaks (R_0_<1, mean size 38, IQR 18-124) across hypothetical emerging viruses spanning a range of wastewater shedding amounts relative to SARS-CoV-2, and proportions of clinically symptomatic infections. To model wastewater detection probability, we first computed the effective number of individuals shedding into wastewater each day, relative to a single SARS-CoV-2 infection on their peak shedding day^19^ (see Methods). We translated daily effective shedders to wastewater detection probabilities, using models fit to SARS-CoV-2 data from New Zealand^53^, and modelled clinical ascertainment by combining the probability of developing clinical symptoms with the probability of diagnosis given symptoms. These simulations allowed us to identify combinations of viral characteristics for which wastewater surveillance added value to conventional clinical surveillance, through unique or earlier outbreak detection.

In general, we would expect wastewater surveillance to provide greatest value for pathogens with high levels of viral shedding in excreta and low rates of clinically symptomatic infection. Simulations showed a threshold relationship between viral shedding amounts and value added, with the position of the threshold strongly influenced by probability of diagnosis given symptoms. We based diagnosis probability on estimates that 10% of ebolavirus spillover infections are detected clinically^54^, and the observation that most emerging virus infections have lower rates of severe and specific symptoms that prompt healthcare seeking and diagnosis than ebolaviruses. Under baseline assumptions (weekly wastewater sampling, 1% of symptomatic infections clinically diagnosed), wastewater surveillance added value when shedding levels exceeded approximately 10^-2^ times that of SARS-CoV-2. The threshold shifted lower (∼10^-4^-10^-3^) when clinical ascertainment was lower (<0.1%) and higher (>10^1^) when ascertainment was greater (<10%) (Figure 2A).

**Figure 2.**
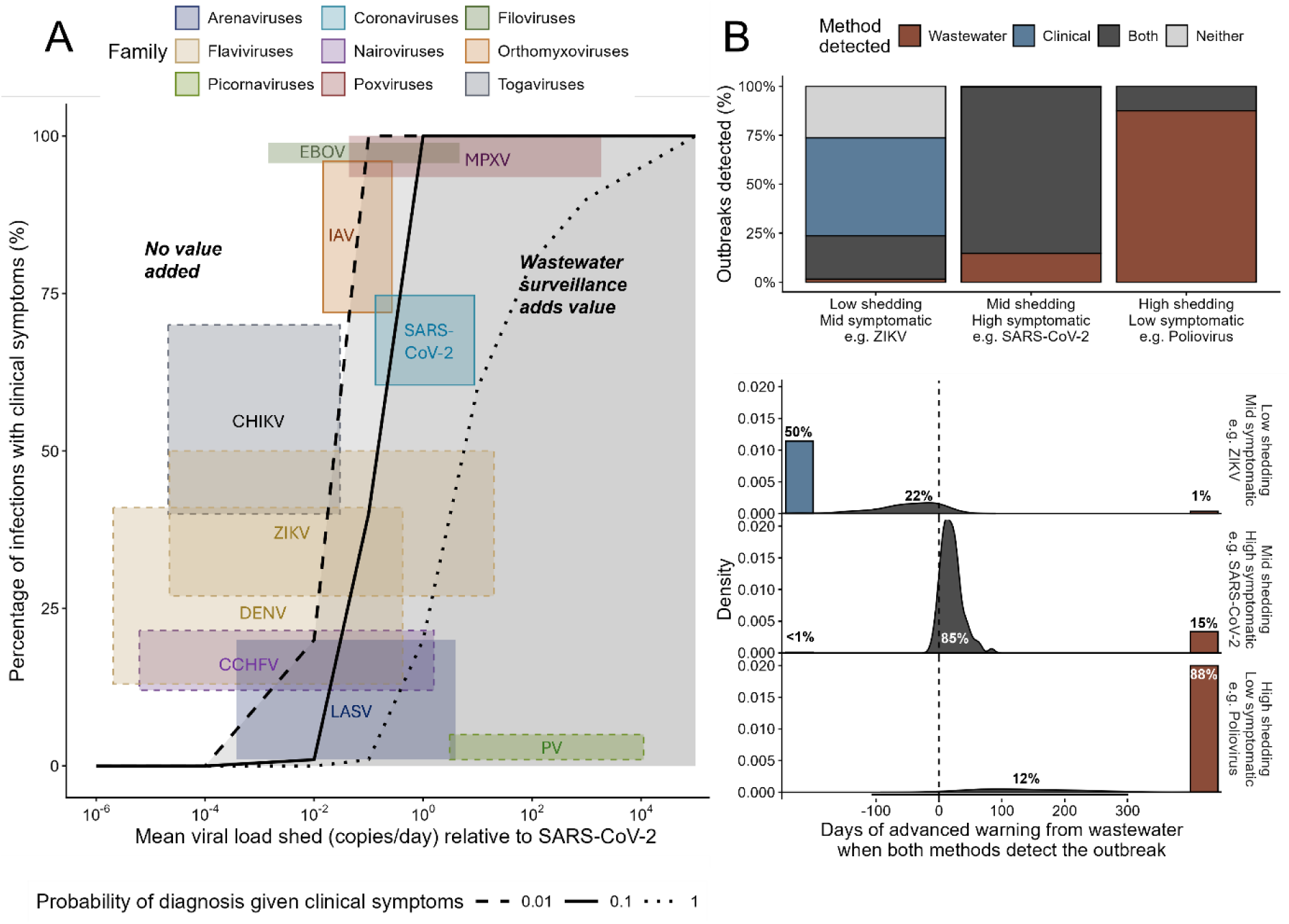
The effect of viral characteristics on the public health value added by wastewater surveillance. **A**. The dependence of the value added by wastewater surveillance on the mean viral load shed to wastewater relative to SARS-CoV-2 (x-axis) and the percentage of infections that result in clinical symptoms (y-axis), under different probability of diagnosis given symptoms (line type: solid = 1, dashed = 0.1, dotted = 0.01).Shaded regions to the right of each line indicate parameter combinations where wastewater increases the proportion of outbreaks detected and/or reduces time to detection. Coloured rectangles, show the approximate parameter space occupied by priority viruses, based on shedding data and the reported proportion of infections that are symptomatic. Solid outlines indicate shedding estimates from comprehensive meta-analyses; dashed outlines indicate estimates based on ≥5 individuals and no outline indicates values based on <5 individuals. Viruses were excluded if shedding data were limited to convalescent or vaccinated shedders, or if variability/uncertainty was not reported. **B.** Proportion of outbreaks detected by clinical surveillance alone (light blue), wastewater surveillance alone (mid blue), both (dark blue) or neither (grey), across 1000 simulated spillovers (∼250 outbreaks >10 infections), assuming 1% probability of diagnosing symptomatic infections. Three parameter combinations approximating central estimates for ZIKV, SARS-CoV-2 and PV are shown. For outbreaks detected by both systems, the lower panel shows the distribution of days of advanced warning provided by wastewater surveillance (negative values indicate earlier detection by clinical surveillance).

To contextualise these shedding thresholds, we tentatively mapped the priority viruses with available viral load estimates onto the parameter space by multiplying the estimates per unit sample volume by average stool and urine volumes produced daily and summing total load shed across urine and feces^44,55^, before accounting for duration and normalising total viral load shed relative to SARS-CoV-2 (Figure 2A). Duration of shedding was highly uncertain for some pathogens (Supplementary material Section 3) but results are far less sensitive to this than to the average viral load estimates which varies across pathogens by orders of magnitude. Wastewater surveillance was most likely to add value for MPXV and SARS-CoV-2-like viruses (both targets of existing successful wastewater surveillance programs), followed by LASV, IAV and EBOV. As an example, for a SARS-CoV-2-like virus, 15% of simulated outbreaks were detected by wastewater surveillance alone, and in the remainder of outbreaks detected by both clinical and wastewater surveillance, wastewater detection provided an average lead time of 20 days (Figure 2B). In contrast, for a ZIKV-like virus, 50% of outbreaks were detected by clinical surveillance alone, just 1% of outbreaks were detected exclusively by wastewater, and in the 22% of outbreaks detected by both systems, clinical detection occurred earlier on average than wastewater detection. Poliovirus (PV), which we included as a benchmark given its history as a successful wastewater target, occupied the region of the parameter space where wastewater surveillance is expected to add the greatest value beyond clinical surveillance. Wastewater alone detected 88% of outbreaks of polio-like virus, with the remaining 12% detected by both wastewater and clinical surveillance, with an average lead time of 173 days in wastewater (Figure 2B).

#### The effect of transmission dynamics

In addition to the viral characteristics that determine within-host dynamics, such as viral shedding and proportion of symptomatic infections, we examined how transmission dynamics could influence the potential value of wastewater surveillance. Holding the amount shed, temporal shedding profile, and symptomatic proportions constant, we varied transmission dynamics across three different zoonotic archetypes 1) sustained seasonal spillover with limited human-to-human transmission (analogous to Lassa virus, Figure 3), 2) a discrete pulse of spillover associated with an animal epizootic with no human-to-human transmission (analogous to Nipah virus in Malaysia), and 3) a short, intense period of elevated spillover risk followed by considerable human-to-human transmission with R_0_ approaching 1, (analogous to some Ebola virus outbreaks) (Figure 4A). The distribution of annual numbers of infections associated with these scenarios can be seen in Figure S1.

**Figure 3.**
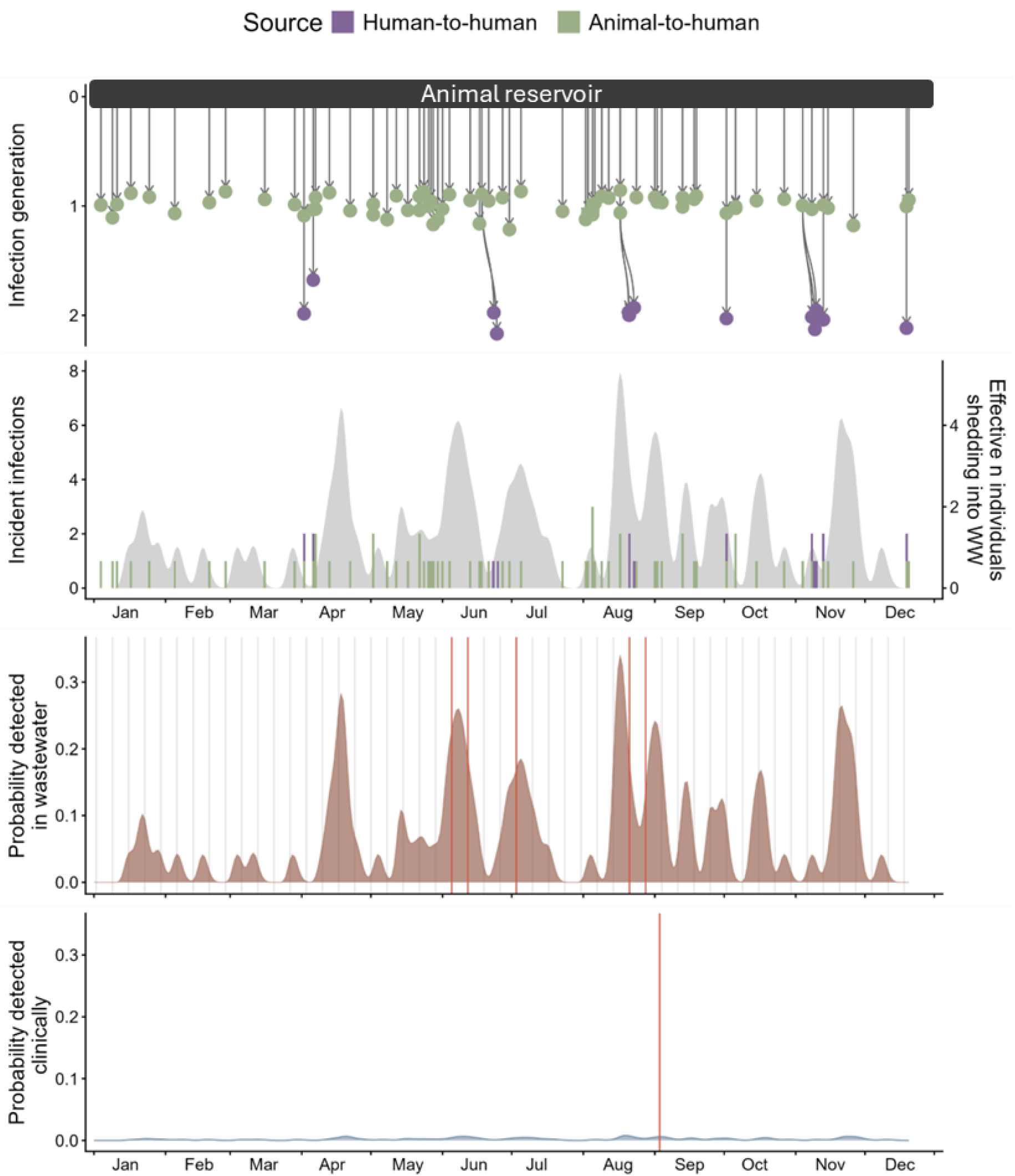
An example of a single stochastic model realisation for transmission of a novel pathogen. This example occurs in a catchment area of 100,000 with a high seasonal spillover rate and low human-to-human transmissibility (e.g. zoonotic archetype 1 Lassa virus-like transmission), with faecal shedding and symptom severity similar to SARS-CoV-2 in 2020, and a 1% chance of diagnosis given healthcare seeking. In the upper panel individual human infections resulting from zoonotic spillover are shown as green points with secondary cases resulting from human-to-human transmission shown in purple and arrows representing transmission. The resultant daily incident infections are shown as a stacked bar chart below with the grey shaded curve indicating the effective number of shedders accounting for the shedding profile over time since infection. The effective number of shedders (see Methods for detailed definition) is translated using models fitted to SARS-CoV-2 data into the probability of wastewater detection shown in brown. The probability of clinical detection based on probability of clinical symptoms, healthcare seeking, diagnosis and probabilistic delays is shown in blue. Grey vertical lines show days when wastewater samples were taken, while red vertical lines indicate detections in this single stochastic realisation of the model.

**Figure 4.**
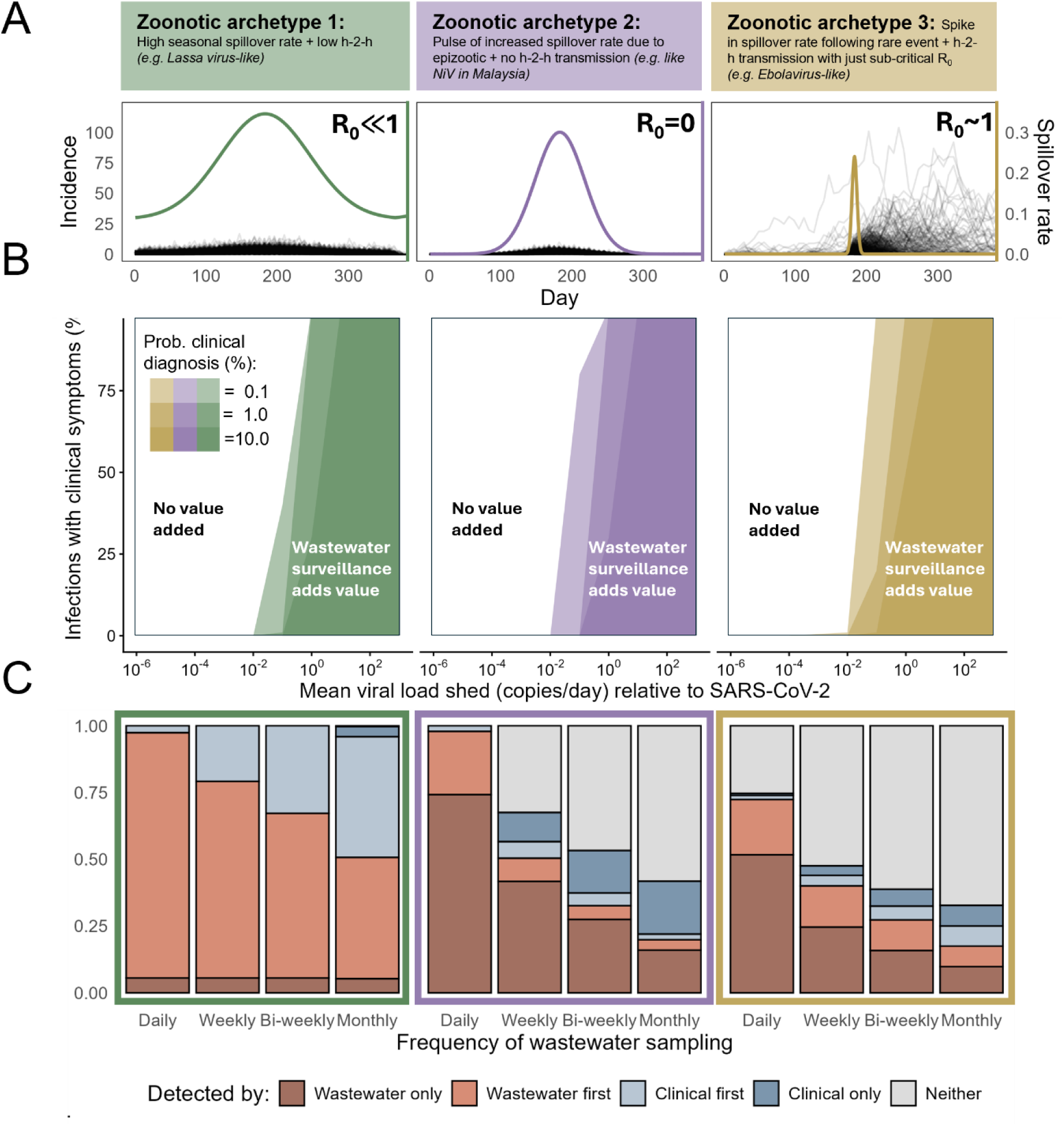
The effect of transmission dynamics on the public health value added by wastewater surveillance. **A.** Three alternative zoonotic archetypes illustrated by their spillover rate shown as a coloured curve over a three year period and annotated with the R0 in humans. Incidence of infections in humans over time, accounting for spillover plus any human-to-human transmission, is shown in black for 100 stochastic model runs **B**. The parameter space in which wastewater surveillance is expected to add value either by detecting otherwise undetected outbreaks or by earlier detection, for each of the three transmission scenarios (shaded = value is added). The darkness of the shading corresponds to the probability of diagnosis. **C**. The proportion of model realisations in which transmission was detected by: wastewater surveillance only; wastewater before clinical; clinical before wastewater; clinical only; or neither surveillance system, for the three archetypes. Results are shown by frequency of sampling, assuming a 1% probability of clinical diagnosis.

The mean viral load shed to wastewater, relative to SARS-CoV-2, required for wastewater surveillance to add value was similar across these three zoonotic archetypes, but shifted slightly depending on the temporal distribution of infections. When cases were distributed more evenly over time, as with sustained spillover in archetype 1, slightly higher shedding levels were generally required compared to archetypes 2 and 3. In contrast, when infections were concentrated in shorter more intense periods of transmission in archetypes 2 and 3 resulting in a greater daily amount of virus shed, wastewater surveillance provided additional value across a broader range of viral shedding amounts and symptomatic proportions (Figure 4B). The type of benefit provided by wastewater surveillance also differed with transmission dynamics. Using SARS-CoV-2 like shedding and clinical delays (Table S6), we compared the contribution of wastewater surveillance across the three zoonotic archetypes (Figure 4C). Under the sustained spillover archetype (archetype 1), wastewater surveillance primarily provided value through earlier detection of infections that would be eventually identified clinically, particularly when the probability of clinical diagnosis was greater than 0.1%. In contrast, in archetypes characterised by more concentrated outbreaks (archetypes 2 and 3), wastewater surveillance more frequently contributed unique detections, identifying transmission that remained undetected by clinical surveillance over a 3-year simulation period. Across all archetypes, the relative contribution of wastewater surveillance decreases as the probability of clinical diagnosis given symptoms increases (rows in Figure 3C). However, for transmission archetype 1, wastewater surveillance continues to add value by frequently enabling earlier detection. For archetypes 2 and 3, in which more transmission events consist of small stochastic outbreaks that are missed by clinical surveillance, wastewater surveillance continues to add value by uniquely identifying transmission, despite higher probabilities of clinical diagnosis.

To assess how sensitive the effects of transmission dynamics on the value added by wastewater were to variation in the temporal distribution of shedding over time, we reran model simulations using a range of alternative shedding profiles. Overall, the value added by wastewater surveillance were relatively robust to changes in profile (Figure S2), which included profiles with more temporally concentrated (spiky) or dispersed (flat) shedding patterns and those with earlier or later peaks in viral load over the course of infection.

#### The effect of frequency of wastewater sampling

More frequent sampling increased both the probability and timeliness of detecting transmission compared with less frequent sampling (Figure 4C). However, lower-frequency sampling still provided substantial value across zoonotic archetypes, for a virus with SARS-CoV-2-like shedding and clinical characteristics. The magnitude and nature of this value varied depended on transmission dynamics. Under sustained spillover with limited onward transmission (archetype 1), increasing sampling frequency primarily enabled earlier detection of infections that would later be detected clinically. In contrast, in archetypes 2 and 3 where infections occurred over a shorter timeframe, more frequent sampling increased the proportion of simulations in which wastewater surveillance uniquely detected transmission missed by clinical surveillance by improving capture of short-lived transmission events. When we varied the probability of clinical diagnosis given symptoms, sampling weekly and even bi-weekly (every 2 weeks) continued to provide earlier or unique detection when the probability of clinical diagnosis was 0.1% or 1% (Figure S3). At higher probabilities of clinical diagnosis, value added by wastewater surveillance declined more rapidly as sampling frequency decreased.

## Discussion

Although wastewater surveillance has been proven effective for tracking enteric viruses (poliovirus), and widespread emerging pathogens (SARS-CoV-2 and MPXV), its broader utility for detecting future viral threats remains underexplored. In this study, we synthesised existing evidence that showed numerous priority viruses from families with pandemic potential, which have not previously been wastewater surveillance targets, are shed in human urine and stool. By combining shedding data with models of transmission and wastewater sensitivity in a quantitative framework, we estimate that many priority viruses have the potential to be detected in wastewater. Wastewater detection of repeat spillover and outbreaks missed by clinical surveillance would inform estimates of pandemic risk and risk-reducing interventions, whilst earlier outbreak detection would afford faster and potentially more successful responses, adding public health value. However, we estimated that this public health value varied substantially across targets based on viral load shed, clinical ascertainment, and transmission dynamics.

Of the viral characteristics we explored in our simulations, viral load shed relative to SARS-CoV-2 was the strongest determinant of the value added by wastewater surveillance. The threshold above which wastewater surveillance would add value ranged from 1/100 to 10 times the viral load shed by SARS-CoV-2, depending on the probability of diagnosis given symptoms. For ebolaviruses infections – the majority of which cause severe disease – it is estimated that 10% of spillover infections are detected clinically^54^, suggesting that the probability of clinical diagnosis could be much lower for novel or emerging diseases with non-specific or moderate symptoms. In such cases, wastewater surveillance could provide unique and early detection even when shedding substantially less than SARS-CoV-2. These relationships between viral load, clinical ascertainment and value of wastewater surveillance have implications for applying wastewater surveillance to emerging viruses, but also other pathogens, including those causing vaccine-preventable diseases.

For many priority viruses it was not possible to reliably estimate mean viral load shed to wastewater. The sensitivity of our findings to virus load estimates highlights the need to better characterise viral shedding in excreta to more reliably assess the value of wastewater surveillance for new targets. This is particularly important for hantaviruses and emerging paramyxoviruses, for which no quantitative data were available. For viruses where viral load entering wastewater could be estimated, we observed substantial variation. Despite uncertainty spanning orders of magnitude, all but MPXV and EBOV were estimated to shed less than SARS-CoV-2.

SARS-CoV and MPXV – both targets of existing successful wastewater surveillance programs^13,56–58^ – had viral shedding amounts and symptomatic proportions consistent with those for which our simulations predict wastewater surveillance would add value for detecting subcritical outbreaks. Despite lower viral load estimates in urine and stool than EBOV, LASV was also among the viruses for which wastewater surveillance was expected to added value, in part due to a lower proportion of symptomatic infections. However LASV shedding amounts and are based on very small sample sizes and more data on shedding in stool is needed to probe this result further. Our results suggest wastewater surveillance is likely to add comparatively less value for detecting subcritical outbreaks of CHIKV, CCHFV, and DENV, than for MPXV, SARS-CoV-2, LASV, EBOV and IAV. However, where clinical surveillance is absent, or limited, wastewater could still add value even for less favourable targets.

Beyond viral characteristics affecting within-host dynamics like viral shedding and clinical presentation, the value of wastewater surveillance is also influenced by transmission dynamics, particularly the distribution of infections through time which differs substantially across viruses owing to spillover and transmission dynamics. Wastewater surveillance added greater value for short, intense outbreaks of human-to-human transmission where clinical detection is more affected by stochasticity, than for sustained low-level spillover. Sampling frequency further shaped this benefit, with more frequent sampling increasing probability and timeliness of detection, particularly for short-lived outbreaks where missed sampling windows could result in outbreaks going undetected. Weekly or fortnightly sampling retained substantial value for pathogens with SARS-CoV-2-like shedding when clinical ascertainment was low, as may be expected for novel pathogens. This is particularly relevant because spillover risk of viruses with pandemic potential is concentrated in low- and middle-income countries, where daily sampling is often infeasible. Identifying when lower-cost lower-frequency wastewater surveillance can still provide meaningful public health value will be important for informing implementation beyond research.

Our study has several limitations arising from data constraints. Firstly, many viral shedding estimates were derived from small sample sizes and infrequent, censored longitudinal sampling, preventing reliable quantification of likely wastewater sensitivity for many priority viruses. Limited reporting of viral loads per unit dry-weight also reduced comparability across studies^20^. Quantitative shedding data largely reflected average viral loads, preventing us from capturing individual heterogeneity or “super shedding”, which has been reported for some viruses^36^. Temporal shedding profiles were unavailable for most priority viruses, meaning we had to use that of SARS-CoV-2 as a baseline and explore several extreme alternatives. However, the estimated value of wastewater surveillance was relatively insensitive to the shedding profile and more strongly influenced by amount shed transmission dynamics.

Secondly, the relationship between daily viral load entering wastewater and probability of detection is based on SARS-CoV-2 data from infected individuals in quarantine facilities in New Zealand, where case and wastewater data were complete. Comparable data are scarce, and the relationship is likely to vary with wastewater system, sample type (e.g. grab vs. trap), assay performance (e.g. different targeted PCR assays or metagenomic sequencing) and any difference in partitioning between solid and liquid fractions. More broadly, anchoring our simulations to this dataset means that our work applies to wastewater surveillance in sewer-based infrastructure, where relatively short travel times mean that recent inputs likely dominate the signal. We assumed shedding contributed to detection only on the day of excretion, which may be less appropriate for non-sewered systems, where longer storage times could make pathogen-specific persistence more important^59^. Nonetheless, qualitative conclusions ranking pathogens by potential value added are likely to be robust across sewered settings.

Finally, generalisability is constrained by the scope of our modelling framework. We assumed exclusively human-derived virus enters wastewater. Contributions from animals could enhance detection sensitivity but complicate source attribution^60^, requiring genetic sequencing and integration with additional data streams^15^. In this context, mutation rate may also influence value added by wastewater surveillance, shaping ability to infer virus origins. We also defined the public health value of wastewater surveillance as detecting otherwise undetected sub-critical outbreaks or providing earlier detection than clinical surveillance. In practice, wastewater surveillance can provide broader benefits, including enhancing situational awareness for widespread pathogens, monitoring viral evolution, and estimating epidemiological quantities e.g. the effective reproduction number (R_t_). For example, wastewater surveillance has demonstrated value for seasonal influenza A, despite this pathogen falling near the boundary of the parameter space where our analysis predicts unique detections and early warning of small outbreaks^61,62^. Establishing an empirical evidence base to validate these findings will be critical to assess when wastewater surveillance for viruses with pandemic potential provides public health value.

In summary, we find that wastewater surveillance holds promise as a complementary tool to address gaps in clinical surveillance for detecting emerging pathogens and understanding spillover dynamics. Our study quantifies how the public health value of wastewater surveillance depends on viral load shed, and how much this varies within and between viral families, emphasising the need to better characterise shedding of priority viruses in human excreta. We expect this modelling framework can support more targeted, evidence-based deployment of wastewater-based epidemiology as it expands to new and emerging threats.

## Methods

### Estimating the total viral load shed to wastewater for priority viruses

To estimate the viral load shed to wastewater, between April and June 2025 we searched the peer reviewed literature available on MEDLINE and EMBASE for studies reporting detection of viral genetic material in urine or stool for each of the priority viruses from the eleven viral families deemed by WHO to pose a high risk of causing a PHEIC. This included *Coronaviridae* (SARS-CoV-2 and MERS-CoV), *Orthomyxoviridae* (Influenza A virus - IAV), *Paramyxoviridae* (Nipah virus - NiV), *Filoviridae* (Zaire ebolavirus - EBOV, Sudan virus - SUDV, Marburg virus - MARV), *Arenaviridae* (Lassa virus - LASV), *Flaviviridae* (dengue virus - DENV, Zika virus - ZIKV, yellow fever virus - YFV), *Nairoviridae* (Crimean Congo haemorrhagic fever virus - CCHFV), *Phenuiviridae* (Rift Valley fever virus - RVFV, severe fever with thrombocytopenia syndrome virus - SFTSV), *Hantaviridae* (Hantaan virus - HTNV, sin nombre virus - SNV), *Poxviridae* (Mpox virus - MPXV) and *Togaviridae* (chikungunya virus - CHIKV). Several key studies characterising the shedding profile of SARS-CoV-2 are available^18–20^. For other pathogens, if a systematic review or meta-analysis of viral load shed in stool or urine was available, this was used as a singular data source for that pathogen, otherwise a comprehensive search was conducted and the results of individual studies were extracted. The data extracted from each study is available in Table S1 along with a more detailed description of our core search strategy including search terms and dates in Table S2.

We extracted quantities including the viral load measured in copies per unit of sample, the percentage of individuals or samples that tested positive, and the duration of shedding, as well as contextual variables including population type (e.g. acute confirmed cases), and the days post symptom onset that samples were taken. When available, fitted model parameters describing the profile of viral shedding over time were also extracted. When viral load measurements were only quantified in a figure these were extracted and digitized using plotdigitizer^63^. If a study plotted individual viral load measurements only, we computed an overall mean to extract as the central value as well as a range of means across individuals to capture a measure of variability. Studies specified units as either copies/g of stool or copies/ml of stool. Only 1 study specified the volume calculations used to reach the estimate, meaning it is not possible to be sure that all measures in copies/ml are directly comparable. Since the single study that did describe the calculation showed that ml referred to ml of the original stool sample rather than stool suspension^27^, we assumed this was the case for all studies. Since most measures of viral load in stool were given in copies/ml, the few measures given in copies/g were converted to copies/ml for comparison assuming stool density of 1.06 g/ml^64^.

In order to compare the likely quantity of viral material shed into wastewater across priority pathogens, we estimated the daily viral load shed to wastewater by an infected individual (VL) as the sum of the daily viral load shed in urine and in stool, which were in turn estimated as follows:

VL(copies) = mean viral load (copies/mL) * volume of waste produced (mL/day)

The median volume of urine produced per day was assumed to be 1420mL/person/day (ranging from 600-2600mL) and the median volume of stool was calculated assuming a wet weight of 128g produced per person per day (ranging from 72-470g), and a density of 1.06g/ml^44,55^. In the case of ebolavirus, each study reported viral load based on a single patient and so we took the range of means across the individuals and studies as an overall estimate. As well as extracting central values, when available, we also extracted the reported uncertainty or variability. When presenting viral load estimates uncertainty was our gold standard. If uncertainty was not available, we presented variability, and if no variability or uncertainty of the central estimate was reported then we use bounds 100-fold above and below the central estimate of the viral load as a conservative default when plotting values, to avoid communicating a false sense of certainty by plotting point estimates.

To compare total viral load shed to wastewater during an infection to that of SARS-CoV-2, and enable translation of simulated number of shedders to probability of wastewater detection, we multiplied the mean daily viral load shed by estimated mean duration of shedding. In some cases the duration was very uncertain but estimates were much less sensitive to the uncertainty in duration compared to in viral load shed which varied over orders of magnitude. In the absence of reliable data on distribution of priority virus shedding across individuals, we assumed that all infected individuals shed during acute infection, and that differences in shedding between individuals and over the course of infection are captured in the mean viral load shed. Details of assumptions used in the estimation of total viral load shed by particular pathogens are given in Table S5.

We based our estimate of the shedding profile, on empirical estimates from Wannigama et al. Of the three key studies^18–20^ characterising SARS-CoV-2 faecal shedding, this study had the greatest sample size, was not limited to severe hospitalised cases, had good support across the duration of infection and reported viral load by wet-weight. In general faecal viral load measures using dry-weight would be preferable, but since there were no other studies that had taken this approach we used wet-weight estimates in an attempt to improve comparability. Measures were excluded for variants BA.2.86 and JN.1, as these demonstrated much higher faecal shedding^19^ and were therefore not representative of strains circulating at the time of collection of the data that we used to estimate the relationship between SARS-CoV-2 infections and wastewater detection^53^.

### Modelling transmission and detection of viruses with pandemic potential

We developed a stochastic modelling framework to simulate spillover, onward human-to-human transmission, and subsequent probabilistic detection by wastewater and clinical surveillance systems (Figure 4).

#### Modelling spillover and transmission

We simulated stochastic daily spillover counts as a Poisson process with a time-varying rate of zoonotic transmission to humans. The time-varying spillover rates were modelled using a circular Gaussian forcing function defined over a period length *P* days (e.g. 365 days to capture annual seasonality), where the circular distance between day *t* and the day the rate peaks, *t_max_*, was defined as

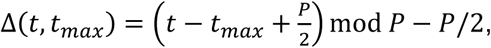

with the spillover rate on day *t* defined as

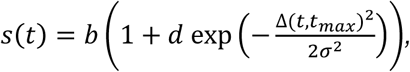

where *b* is the baseline spillover rate, *d* controls the magnitude of the peak relative to baseline and *σ* determines the spread of the peak over time. The number of human infections resulting from spillover on day *t* day is then given as

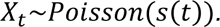

These human infections were then used to seed onward human-to-human transmission which we modelled using a stochastic branching process. For each infected individual *i*, the number of secondary infections *Z_i_* was drawn from an offspring distribution with a mean corresponding to the reproduction number *R_0_*. The number of secondary cases was assumed to be negative binomially distributed to capture overdispersion:

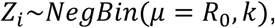

where *µ* is the mean number of secondary cases and *k* is the overdispersion parameter controlling the heterogeneity in transmission. Superspreading dynamics are represented by small *k* values.

Since we were exclusively modelling the subcritical human-to-human transmission characteristic of early stuttering pre-pandemic chains resulting in very small numbers of cases we did not adjust R_t_ based on depletion of susceptible individuals in our core analysis as this would be negligible in a large catchment area of 100,000. For each simulated infection, epidemiological and clinical characteristics were assigned probabilistically, with generation times, incubation periods, and other delays drawn from gamma distributions. These were mainly based on SARS-CoV-2 when simulating outbreak scenarios (Table S6).

#### Modelling wastewater surveillance

We combined the time series of incident infections with a viral shedding profile to generate a time series of the effective number of shedders per day for different scenarios. Since the shedding profile in urine or stool has not been well characterised for any priority viruses with pandemic potential, we used the shape of the faecal shedding profile of SARS-CoV-2^19^ as a proxy and scaled the profile so that the total viral load shed (AOC) varied between 10^-2^ -10^6^ fold relative to SARS-CoV-2 to cover the range of total viral load shed across priority pathogens estimated from mean viral load measurements, average daily volumes of excreta produced, and durations of shedding found in the literature (Figure 1). As a sensitivity analysis, we also repeated our simulations using spikier profiles that distributed the majority of viral load shed over a shorter duration, and flatter profile that distributed the viral load more evenly over the duration of shedding. On the day that shedding peaked post infection, a person with SARS-CoV-2 was counted as 1 effective shedder, and proportionally less on the days before and afterwards according to the shedding profile. Similarly, if we were modelling detection of a virus that shed 10x less than SARS-CoV-2, an individual infected in this case would count as 0.1 effective shedders on their peak day of shedding, and days either side of this would be scaled accordingly. Owing to waste travel times in sewered systems, and half-lives of enveloped RNA viruses and viral material, recent inputs are expected to dominate measured signals in wastewater. We therefore assume that viral shedding contributes to detection on the day that it is shed only. We do not consider potential differences in in rates of decay between pathogens over the time frame of one day.

In order to translate the effective number of shedders into a probability of wastewater detection, we fitted a wastewater detection sensitivity model to a dataset with likely complete infection ascertainment from a large COVID-19 quarantine facility in New Zealand^53^. Published models of sensitivity based on these data were fitted prior to detailed characterisation of SARS-CoV-2 faecal shedding dynamics and therefore used a nasal shedding proxy to translate incidence into effective shedders. They also employed a standard logistic formulation which, given the limited number of observations with very low case numbers, resulted in the left tail of the predicted detection probabilities being relatively unconstrained and estimated a 20% probability of detection in the absence of any cases. We therefore refit a model to the wastewater and case data^53^, using an empirically derived faecal shedding profile estimate effective shedders and adopted a log_10_ transformed logistic functional form and a Hill function both constrained to pass through the origin, such that zero infections corresponded to zero probability of detection at the WWTP (Table S7, Figure S4). The model was fit using Hamiltonian Monte Carlo (HMC) implemented in rstan^65,66^. Uninformative normal priors were applied to all model parameters. Model fit was compared using the loo R package version 2.9.0^67,68^. Using the faecal shedding profile to determine effective case numbers afforded a better fit than the nasal profile (Table S7). Constraining the model to pass through the origin led to a small but not meaningful decrease in fit^69^, and so the log_10_ transformed logistic regression model that passed through the origin was used in our core analysis. Although we do not escape the issue of relatively uninformed probabilities at low case numbers, it was particularly important for our purposes to ensure that detection probabilities approached zero at low prevalence as our scenarios of interest focuses on spillover and early transmission of viruses including those which may be shed at substantially lower levels than SARS-CoV-2. Using the existing standard logistic model directly would have led to over estimation of the utility of wastewater surveillance in such scenarios.

Stochastic wastewater detection events are drawn from a binomial distribution on the days in which sampling is simulated (e.g. every 7^th^ day for weekly sampling in the example in Figure 3) using the probability determined using the number of effective shedders contributing to the wastewater on that day, based on the log_10_ transformed logistic regression model.

#### Modelling clinical surveillance

Clinical detection was modelled based on the successful diagnosis of at least 1 case, which required sequential probabilistic events: the individual becoming symptomatic, accessing healthcare and being accurately diagnosed. For known viruses of interest, the probability of an infection becoming symptomatic was parameterised using the proportion symptomatic reported in the peer-reviewed literature or on public health agency websites (Table S8). For the purposes of presenting the probability of clinical detection over time in Figure 3, we combined the infection incidence time series with the per-infection probability of clinical detection *q* at lag *k* days defined as:

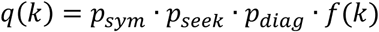

where *p_sym_* is the probability of developing symptoms, *p_seek_* is the probability of seeking and accessing healthcare given symptoms, *p_diag_* is the probability of diagnosis conditioned on healthcare access, and *f(k)* is the convolution of the delays from infection to symptom onset, onset to healthcare access, healthcare access to diagnosis. For each day *t*, we then calculated the probability that at least one clinical detection arises from all prior infections as:

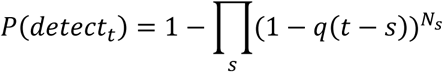

Where *N_s_* is the number of infections occurring on day *s* (prior to day *t*).For *p_diag_*, we assumed that 1% of symptomatic infections would be diagnosed, and varied this from 0.1% to 10% as a sensitivity analysis. This is based on estimates that 10% of ebolavirus spillover infections are detected clinically^54^, and the observation that most emerging virus infections have lower rates of severe and specific symptoms that prompt healthcare seeking and diagnosis than ebolaviruses.

Performance of clinical and wastewater surveillance were compared in terms of unique detections and, in the case that both systems detected an outbreak, time from the first infection to first detection. When assessing the effect of temporal transmission dynamics, the number of infections rather than days prior to detection was used, though there was not a meaningful difference in results between the two metrics.

The model framework is implemented in R version 4.5.0^70^.

## Supporting information

Table S1 data dictionary

## Data Availability

All data produced in the present study are available within the manuscript itself, the supplementary material and associated public github repository.

https://github.com/waspp-consortium/wastewater_sim

## Acknowledgements

We thank Dr Joanne Hewitt and Bridget Armstrong for sharing extra information regarding the dataset behind their publication on wastewater detection and SARS-CoV-2 cases in a COVID-19 quarantine facility in New Zealand. We thank the members of the Wastewater Surveillance for Pandemic Prevention (WaSPP) consortium, the Vaccine Epidemiology Research group, and Dr Elana Chan for insightful discussions that improved this work. This study was funded by the Gates Foundation (grant INV-076271). All authors also acknowledge funding from the MRC Centre for Global Infectious Disease Analysis (reference MR/X020258/1), funded by the UK Medical Research Council (MRC). This UK funded award is carried out in the frame of the Global Health EDCTP3 Joint Undertaking.

## Data and code availability

The framework developed and presented here is available as an R package wastewatchR release v0.0.2 (tag: paper1_version1) available from https://github.com/mrc-ide/wastewatchR. The further analysis code and data generated by this study are available at https://github.com/waspp-consortium/wastewater_sim.

