## Supplementary Material for "The public health value of wastewater surveillance for viruses with pandemic potential: a modelling study"

### Section 1. Review results

For Table S1 containing the complete extracted dataset that resulted from the review please see the separate .xlsx file.

### Section 2. Search strategy

We first searched PubMed for systematic reviews and meta-analysis of shedding of viruses with pandemic potential in urine and stool. We defined viruses with pandemic potential as those identified by WHO as priority pathogens within the 11 viral families designated as posing a high risk of causing a PHEIC or pandemic in the 2024 pathogen prioritisation exercise^1^. These viruses are *Coronaviridae* (SARS-CoV-2 and MERS-CoV), *Orthomyxoviridae* (Influenza A virus - IAV), *Paramyxoviridae* (Nipah virus - NiV), *Filoviridae* (Zaire ebolavirus - EBOV, Sudan virus - SUDV, Marburg virus - MARV), *Arenaviridae* (Lassa virus - LASV), *Flaviviridae* (dengue virus - DENV, Zika virus - ZIKV, yellow fever virus - YFV), *Nairoviridae* (Crimean Congo haemorrhagic fever virus - CCHFV), *Phenuiviridae* (Rift Valley fever virus - RVFV, severe fever with thrombocytopenia syndrome virus - SFTSV), *Hantaviridae* (Hantaan virus - HTNV, sin nombre virus - SNV), *Poxviridae* (Mpox virus - MPXV) and *Togaviridae* (chikungunya virus - CHIKV). Central estimates of viral load shed in stool and the temporal profile of shedding were extracted from the reviews found.

With the exception of SARS-CoV-2, for which several well-known studies are available characterising shedding profile in stool, if a systematic review or meta-analysis was not available for a given pathogen, a comprehensive search of MEDLINE and EMBASE was conducted using the search terms details in Table S2, on the dates specified. No date restrictions were used except for in the case of MERS-CoV where the search was restricted to 2012 and onwards. The search was conducted by a single individual.

Table S2. Searches conducted to synthesise evidence of shedding of priority viruses in urine and stool.

| Viral family | *Priority virus* (common abbrev.) | Search terms | Date of search |
| --- | --- | --- | --- |
| Arenaviridae | *Mammarenavirus lassaense* (LASV) | (lassa fever or lassa virus) and (shedding or urine or feces or faeces or stool) | 13/05/2025 |
| Coronaviridae | *Subgenus Merbecovirus* (MERS-CoV) | (MERS-CoV or MERSCoV or Middle East Respiratory Syndrome Coronavirus) AND (shedding or urine or stool or faeces or feces) | 28/04/2025 |
|  | *Subgenus Sarbecovirus (SARS-CoVs)* | Full search not conducted for SARS-CoV-2 due to the availability of several well-known studies characterising the shedding profile of SARS-CoV-2, and the volume of literature precluding a truly comprehensive and timely search. | NA |
| Filoviridae | *Orthoebolavirus*  *Zairense* (EBOV) | (ebola virus or ebolavirus) and (urine or faeces or feces or stool or stool or shedding) | 16/05/2025 |
|  | *Orthoebolavirus sudanense* (SUDV) | (ebola virus or ebolavirus or sudan virus) and (urine or faeces or feces or stool or stool or shedding) | 16/05/2025 |
|  | *Orthomarburgvirus marburgense* (MARV) | marburg virus and (urine or faeces or feces or stool or stool or shedding) | 18/06/2025 |
| Flaviviridae | *Orthoflavivirus zikaense* (ZIKV) | zika virus and (rna or shedding) and (urine or faeces or feces or stool or shedding) | 28/06/2025 |
|  | *Orthoflavivirus dengue* (DENV) | dengue virus and (rna or shedding) and (urine or faeces or feces or stool) | 28/06/2025 |
|  | *Orthoflavivirus flavi* (YFV) | yellow fever virus and (rna or shedding) and (urine or faeces or feces or stool) | 28/06/2025 |
| Hantaviridae | *Orthohantavirus sinnombreense* (SNV) | (hantavirus or hanta virus or sin nombre virus or four corners virus or muerto canyon virus or convict creek virus) and (shedding or rna) and (urine or faeces or feces or stool) | 28/04/2025 |
|  | *Orthohantavirus hantanense* (HTNV) | (hantavirus or hanta virus or hantaan virus) and (shedding or rna) and (urine or faeces or feces or stool) | 28/04/2025 |
| Nairoviridae | *Orthonairovirus haemorrhagiae* (CCHFV) | (crimean congo hemorrhagic fever or crimean conge haemorrhagic fever or cchf) and (shedding or urine or stool or faeces or feces) | 13/05/2025 |
| Orthomyxoviridae | *Alphainfluenzavirus Influenzae H1, H2, H3, H5, H6, H7 and H10.* (IAV) | Systematic review available for Influenza A – full search not conducted | NA |
| Paramyxoviridae | *Henipavirus nipahense* (NiV) | (nipah virus or niv) and (shedding or urine or faeces or feces) | 15/05/2025 |
| Phenuiviridae | *Bandavirus dabieense* (SFTSV) | (severe fever with thrombocytopenia syndrome or SFTS) and shedding or urine or feces or faeces or stool | 14/05/2025 |
|  | *Phlebovirus riftense* (RVFV) | (rift valley fever or rift valley fever virus) and (stool or feces or faeces or urine or shedding or RNA or viral load) | 14/05/2025 |
| Poxviridae | *Orthopoxvirus variola* (VARV) | (smallpox virus or small pox virus or variola virus) and (urine or shedding or feces or faeces or stool) | 19/06/2025 |
|  | *Orthopoxvirus monkeypox* (MPXV) | (mpox or monkeypox or monkey pox) and (urine or shedding or feces or faeces or stool or viral dynamics) | 14/05/2025 |
| Togaviridae | *Alphavirus Venezuelan* (VEEV) | Venezuelan equine encephalitis virus and (shedding or urine or feces or faeces or stool) | 13/05/2025 |
|  | *Alphavirus chikungunya* (CHIKV) | chikungunya virus and (shedding or urine or feces or faeces or stool) | 13/05/2025 |

### Section 3. The temporal shedding dynamics of priority viruses

Table S3: Reported viral shedding profiles of priority viruses in urine or stool

| Virus* | Study | Sample type | N indiv. | Days PSO** | Temporal profile type | Summary |
| --- | --- | --- | --- | --- | --- | --- |
| SARS-CoV-2 | Hoffman et al 2023^2^ | Stool | 27 | 6 to 30 | Viral concentration (copies/ml) | Bayesian hierarchical generalised gamma model with an exponential temporal decay function fitted to viral concentrations from stool from hospitalised patients. Authors estimated the half-life of 34 hours, implying >90% of shedding occurs in the first week. Note limited data prior to 6 days PSO means this shedding estimate in the first week is largely unconstrained. Concentrations dip below limit of detection at ~20 days based on our visual assessment of Figure 2 of the study. |
|  | Wannigama et al 2024^3^ | Stool | 113 | -3 to 21 | Viral concentration (copies/mg) | Distribution of viral concentrations in stool samples from community SARS-CoV-2 infections among vaccinated individuals, plotted by day PSO showing shedding peaks around 9 days PSO and tapers towards the limit of detection at 21 days PSO. |
|  | Arts et al 2023^4^ | Stool | 48 | -3 to 28 | Viral concentration (copies/mg-dry weight) | A model was not fitted but the raw data are plotted over time along with the proportion of positive samples over time post symptom onset. Temporal trends appear relatively consistent with Wannigama et al. as judged visually. |
| MPXV | Kim et al 2023^5^ | Urine | 21 | 1 to 17 | Viral concentration (Ct) | Fit a local polynomial regression model to the results of a meta-analysis. Authors report the a slope change around 7 days PSO, with Ct values increasing after this point. |

**See Table S2 for full species names, **Post Symptom Onset*

Table S4: Reported estimates of the duration of shedding of priority viruses in urine or stool

| Virus* | Study | Sample type | Duration (days) | Sample size | Method/definition | Days PSO** covered |
| --- | --- | --- | --- | --- | --- | --- |
| DENV | Andries et al 2015^6^ | Urine | 16 | 57 | Last day detected | 1-120 |
|  | Hirayama et al 2012^7^ | Urine | 16 | 53 | Last day detected | 0-33 |
| IAV | Lowry et al 2023^8^ | Stool | 27 | 2 | Last day detected | NA |
| MPXV | Piralla et al 2024^9^ | Urine | 9 | 14 | Kaplan-Meier estimate of median time to clearance | 0-30 |
|  | Cordeiro et al 2024^10^ | Urine | 8 | 14 | Farazdaghi-Harris density function – day after which 90% expected to be undetectable | 0-14 |
| CHIKV | Martins et al 2022^11^ | Urine | 26 | 35 | Median from fitted Weibull | 0-95 |
| ZIKV | Paz-Bailey et al 2017^12^ | Urine | 11 | 136 | Median from fitted Weibull, accounting for interval censoring. 95^th^ percentile was 34 days. | 0->60 |

**See Table S2 for full species names. **days Post Symptom Onset that samples were taken*

We did not find reported quantitative estimates of the duration of shedding of the remaining viruses/sample types not included in Table S3 or S4. However, for LASV, EBOV, CCHFV and ZIKV we could infer information about the rough duration from raw longitudinal data or figures to support assumed windows of duration to inform our estimates of the potential viral load shed to wastewater relative to SARS-CoV-2, as follows:

**LASV**

We did not find reports of any quantitative estimates of the duration of shedding for LASV, but evidence suggests prolonged shedding in urine. The largest study of 37 shedders found some to shed 3-6 months after symptom onset with the majority ceasing between 1-3 months^13^. When estimating total shedding relative to SARS-CoV-2, we assumed a shedding duration of 2-6 weeks in urine to reflect the range of the majority of reported estimates across the four longitudinal studies available^13–17^. We only found a single case study which had looked for LASV in stool^14^. Viral load was measured to be 10-fold higher than that in urine. Since no longitudinal estimates of shedding in stool were available, we assumed a potential range from 1 week up to the upper bound for urine of 6 weeks.

**EBOV**

We did not find any studies that focused on estimating the duration of shedding of EBOV in urine or stool. Records of shedding over time were limited to case studies that each focused on a single patient. We based our assumed range of shedding duration on the range of the last day of shedding extracted from longitudinal plots of viral load in the urine of 6 individuals^18–23^ (1-5 weeks) and in the stool 4 individuals^19,20,23,24^ (1-4 weeks).

**CCHFV**

Although we did not find any studies that reported a quantitative estimate of the duration of shedding of CCHF in urine or stool, we did find several studies which presented viral load estimates over time for small numbers of individuals^25–29^. For urine, 5 studies involving a total of 17 individuals, measured maximum durations of shedding mostly ranging from around 1-2 weeks which we took forward into our assumptions, with one example of more prolonged shedding up to 7 weeks^28^. The single study that looked at shedding in stool suggested a duration of around 1 week based on samples from 6 individuals^27^.

**ZIKV**

A key study involving 136 individuals estimated the median duration of shedding to be 11 days in urine with the 95^th^ percentile at 34 days PSO^12^. Two smaller studies involving a total of 11 individuals took longitudinal measurements of ZIKV shedding in urine suggesting a shedding duration of 1-2 weeks but with considerable right censoring affecting the larger study. We therefore assumed an average shedding duration spanning 1-3 weeks. There is very limited evidence that ZIKV is shed in stool, though it has been found in rectal swabs^30^.

**IAV**

Despite finding reported estimates of IAV shedding duration in stool, we did not find data on the shedding duration of IAV in urine. However, evidence suggested that the majority of individuals have a positive urine sample suggesting IAV is meaningfully shed in urine^8^. One study was identified which sampled urine longitudinally and measured shedding up to three weeks post symptom onset, however observations were limited to a single severely ill individual^31^. We therefore assumed a conservative lower bound of one week and an upper duration of three weeks (close to that in stool) for IAV in urine when estimating likely total shedding contribution to wastewater relative to SARS-CoV-2.

**Table S5**: Virus specific assumptions used in estimates of daily viral load shed relative to SARS-CoV-2

| Virus* | Assumptions |
| --- | --- |
| LASV | No measure of uncertainty or variability in average viral load was available so we applied a conservative estimate of +/-100 times the central estimate.  The duration of shedding in stool was not available but there is evidence that LASV can be present in stool (detected up to day 11, at 10,000 copies/mL which is 10x mean in urine^14^). We therefore assumed a range of shedding durations in stool from 1 week as a conservative lower bound, to the same upper bound as that seen in urine. |
| EBOV | The range of mean viral load shed was taken across individual case studies to represent variability. |
| ZIKV | We assume no shedding contribution from stool |
| DENV | We assume no shedding contribution from stool |
| CCHF | Mean viral load estimated from raw published data from individuals who did not receive ribavirin (as those who did receive treatment shed much less)^27^. |
| MPXV | Viral load has only been quantified in terms of copies/unit for urine, but multiple studies measured Ct values in both stool and urine – one of which made this data available (n=12)^32^. Assuming difference in viral load is 2^ the difference in Ct led us to assume that viral load in stool was 370 fold higher than in urine +/- 100 times this central estimate. |
| CHIKV | We assume no shedding contribution from stool |

**See Table S2 for full species names*

### Section 4. Supplementary model outputs and sensitivity analyses

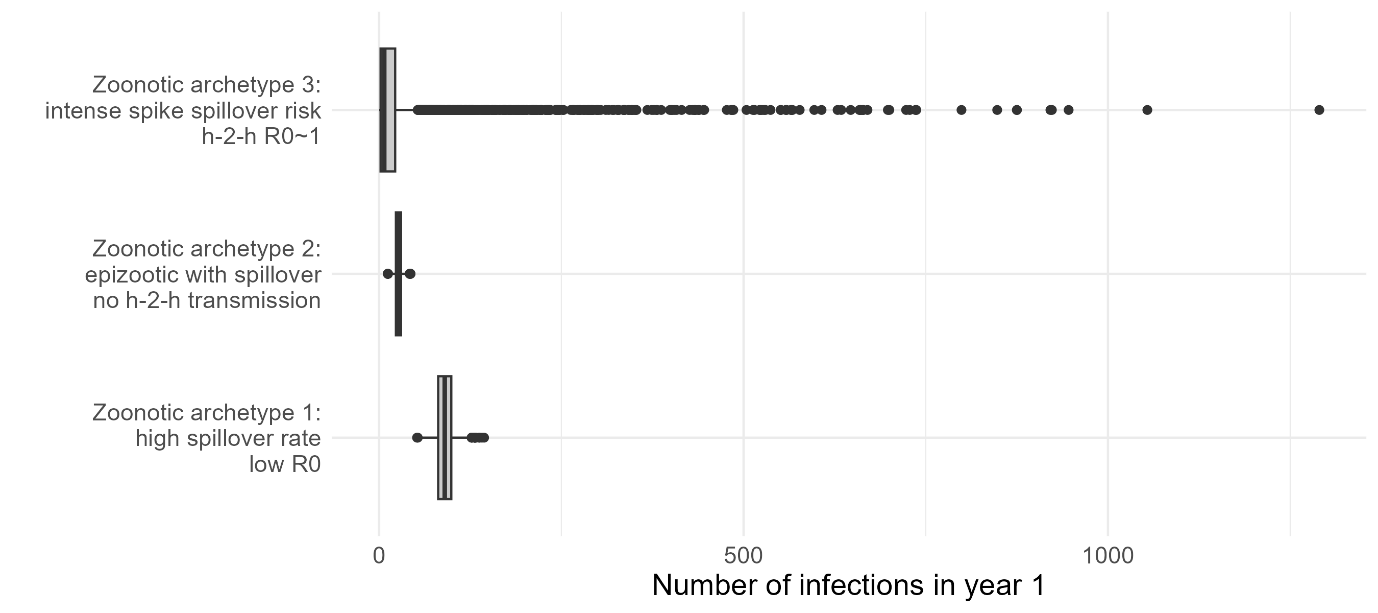

**Figure S1.** The distribution of annual incidence of infections (x axis) across the 3 zoonotic archetypes (y axis). Average infection numbers are similar across archetypes but for zoonotic archetype 3 in which R_0_ approaches 1, we see that in the majority of model runs spillover results in a small number of infections, it is occasionally followed by very large outbreaks.

**Table S6**: Additional epidemiological parameters used in simulations

| Parameter | Values | Rationale |
| --- | --- | --- |
| Generation time | Gamma distribution with shape = 12 and rate = 2 | Mean of 6 days chosen to simulate SARS-CoV-2-like interval^33–35^ |
| Incubation period | Gamma distribution with shape = 5.8 and rate = 0.95 | Based on estimates for wild type SARS-CoV-2^36^ |
| Time from symptom onset to healthcare contact | Gamma distribution with shape = 14 and rate = 2 | Based on estimates for wild type SARS-CoV-2^37^ |
| Time from healthcare contact to diagnosis | Gamma distribution with shape = 6 or 14, and rate = 2 | Mean of 3 or 7 days to reflect either relatively rapid testing and test processing, or 1 week delay |
| Initial population level immunity | 0 | Chosen to reflect emergence of a virus that is novel in the area, meaning very few people will have protective immunity. |
| R0 | 0.99 in baseline outbreak simulations for Figure 2 (or otherwise as stated in the main text for the specific zoonotic archetypes explored). | Chosen to reflect a just sub-critical value leading to self-limited outbreaks with size highly affected by stochastic effects. |
| Temporal shedding distribution | Gamma distribution with mean = 13.1 days, sd = 2.4 days.  Variations for sensitivity analysis, normalised to ensure same total shedding over time:   - spikier (sd*0.25) - flat over 21 days - early peak (mean 4.6 days, sd 3.2 days) | Fitted to SARS-CoV-2 faecal shedding data presented by Wannigama et al^3^. Variants JN.1 and BA.286 were excluded as they had much higher faecal shedding and were not representative of early SARS-CoV-2 strains circulating at the time that the data we used to estimate the relationship between shedding and wastewater sensitivity was collected^38^.  Early peak in sensitivity analyses based on estimated nasal shedding profile of SARS-CoV-2^39^ |

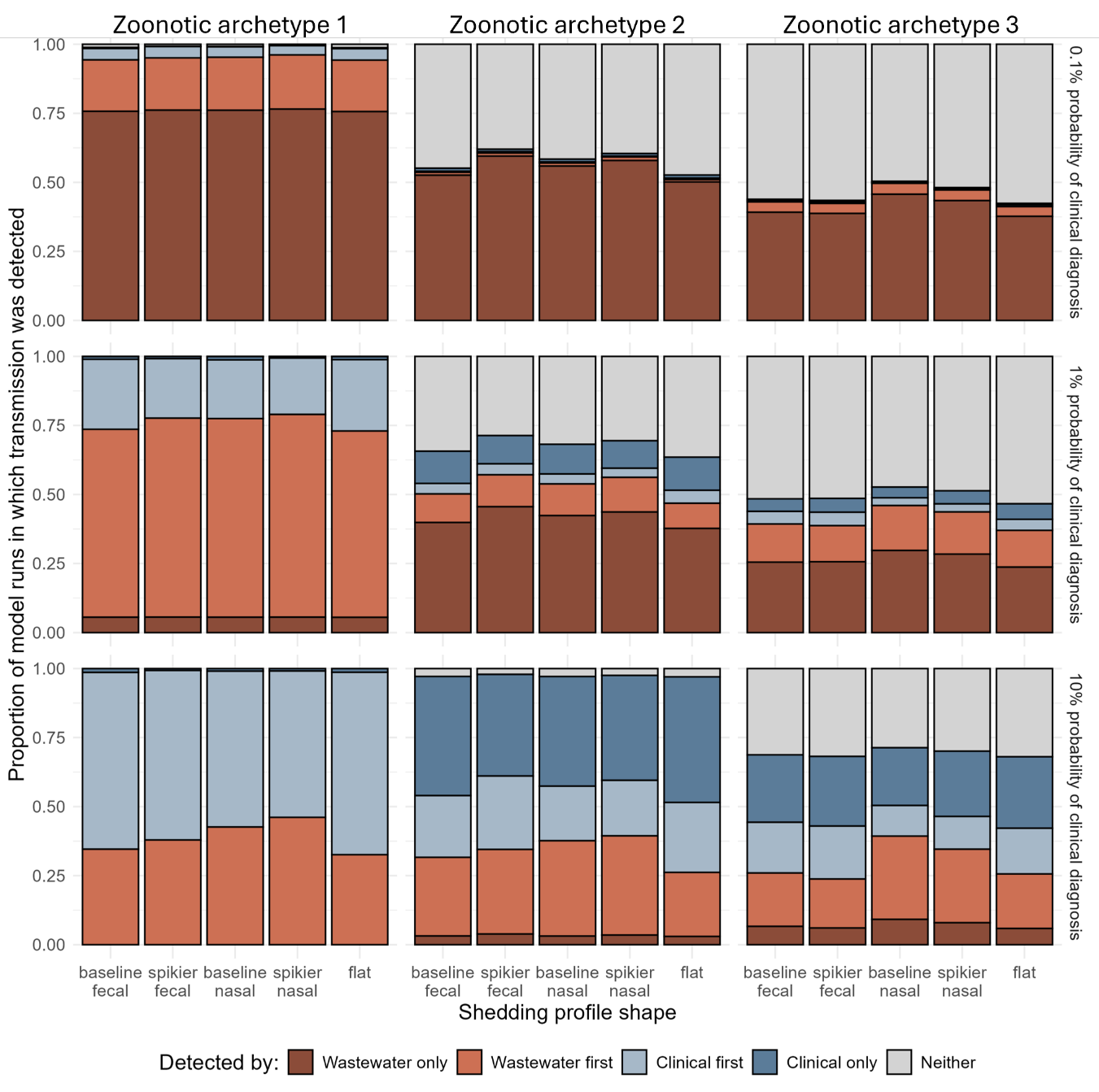

**Figure S2.** The public health value added by wastewater surveillance is robust to changes in shedding profile shape. The breakdown of detections from wastewater, clinical and both are similar across spikier profiles (same total viral load but shed over a shorter time), to flat profiles (more diffuse shedding with equal intensity across the full duration of the shedding period), and from faecal-like profiles which peak longer after infection (based on SARS-CoV-2^3^), to nasal-like profiles which peak soon after infection.

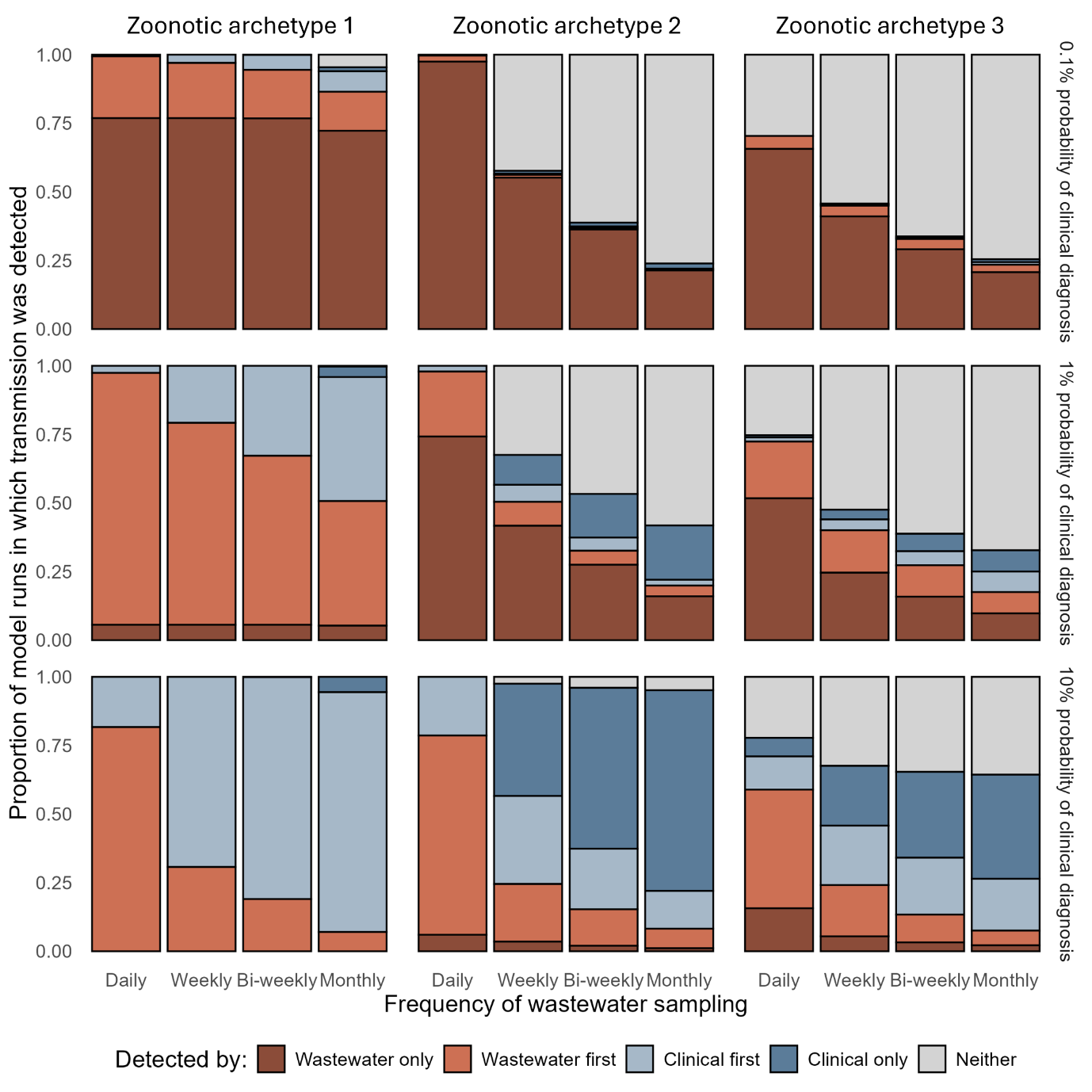

**Figure S3.** The proportion of model realisations in which transmission was detected by: wastewater surveillance only; wastewater before clinical; clinical before wastewater; clinical only; or neither surveillance system shown for alternative probability of clinical diagnosis (rows). Results are shown by zoonotic archetype and frequency of wastewater sampling.

### Section 5. Supplementary methods

**Table S7.** Comparison of model fit represented by the difference in expected log pointwise predictive density (ELPD) when using leave one out cross-validation, across different shedding profiles and model forms. The difference in standard error (SE) is also shown for context when interpreting ELPD.

| Shedding profile | Model | ELPD diff | SE diff |
| --- | --- | --- | --- |
| Fecal | Standard logistic | 0.0 | 0.0 |
|  | Log10-transformed logistic | -2.5 | 1.0 |
|  | Hill function | -2.6 | 1.0 |
| Nasal | Standard logistic | -4.6 | 3.5 |
|  | Log10-transformed logistic | -5.2 | 3.2 |
|  | Hill function | -5.2 | 3.2 |

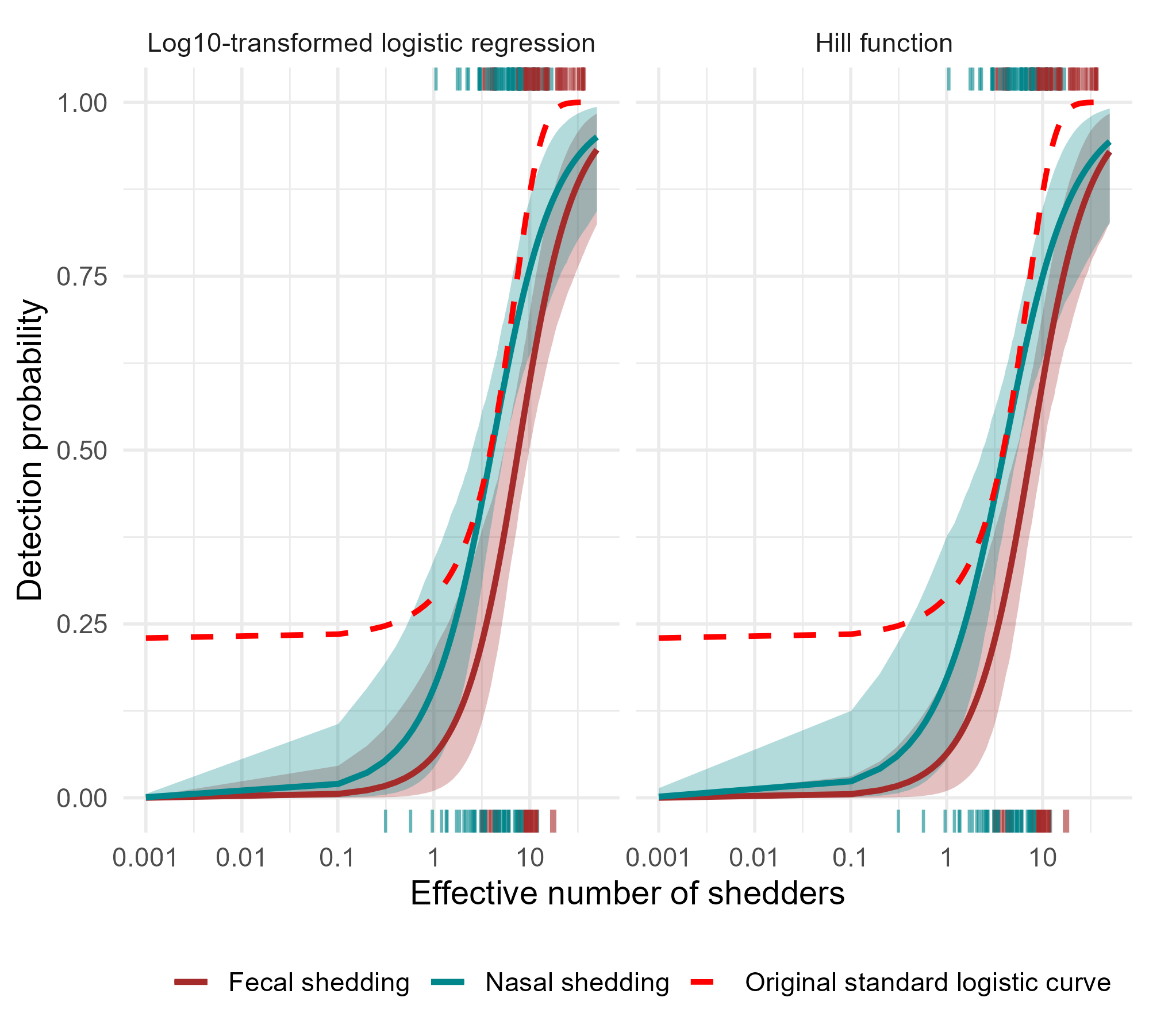

**Figure S4.** The predicted probability of detection in wastewater based on effective number of shedders. Two alternative models (columns) were fitted to 110 observations of effective case numbers and wastewater treatment plant detections. The red dotted line shows the predicted probability of detection based on the published standard logistic regression by Hewitt et. al. for comparison. Lines are coloured according to the underlying shedding profile used. The transparent ribbons represent the 95% credible intervals. The rug plots on the top of the plots show the distribution of effective numbers of shedders on days were detection occurred, and those on the bottom indicate effective numbers of shedders on days where detection did not occur, coloured by the shedding profile used.

**Table S8:** The percentage of infections that develop clinical symptoms for priority viruses used in Figure 2A

| Virus* | Symptomatic infections (%) | Comments | Source |
| --- | --- | --- | --- |
| SARS-CoV-2 | 68 (95%CI: 61-75) | Meta-analysis. Based on omicron | ^40^ |
| LASV | 1-20 | Official websites for WHO, CDC and ECDC report 20% but do not provide data sources. | ^41,42^ |
| EBOV | 98 (95%CI: 96-99) |  | ^43^ |
| ZIKV | 27-50 | Range across 3 locations accounting for imperfect diagnostics | ^44^ |
| DENV | 13-41 | Range across immune states (18% of primary infections, 13% of secondary <1 year and 41% of secondary >1 year). | ^45^ |
| CCHF | 12-22 | Range across studies | ^46,47^ |
| MPXV | 94-100 | Range across studies | ^48^ |
| IAV | 84 (81-87) | Meta-analysis. We used the pooled estimate across outbreaks as this fits the emergence scenario we are modelling better than estimates from longitudinal surveillance of seasonal infleunza-like-illness which estimate 15-35% of infections are symptomatic. | ^49^ |
| CHIKV | 55 (95%CI: 40-70) | Values based on fitted gaussian model of % symptomatic by seroprevalence. We took the values for % symptomatic when seroprevalence is low (between 0-20%) given we are considering emergence. | ^50^ |
| PV | 1-5 | But <1% have paralysis | ^51,52^ |

**See Table S2 for full species names*

41.	World Health Organisation. WHO Lassa fever. https://www.who.int/health-topics/lassa-fever.

42.	Goios, A., Varma, A., Kagia, C., Otiende, M. & Suykerbuyk, P. Report: Enable Lassa Research Programme Mid-term Workshop.

51.	Khan, O. A. & Heymann, D. L. Poliovirus infection - Symptoms, diagnosis and treatment | BMJ Best Practice. https://bestpractice.bmj.com/topics/en-gb/902.

52.	CDC. *CDC Yellow Book 2026: Health Information for International Travel*. (Oxford Academic, 2026).
